# Optimal dose and safety of intravenous favipiravir in hospitalised patients with SARS-CoV-2 infection: a Phase Ib, open-label, dose-escalating, randomised controlled study

**DOI:** 10.1101/2025.06.09.25329141

**Authors:** Tim Rowland, Richard FitzGerald, Elizabeth Challenger, Laura Dickinson, Laura J. Else, Lauren Walker, Colin Hale, Victoria Shaw, Callum Kelly, Rebecca Lyon, Jenn Gibney, Karim Dhamani, Margaret Irwin, Yvanne Enever, Michelle Tetlow, William Wood, Helen Reynolds, Justin Chiong, Orod Osanlou, Henry Pertinez, Katie Bullock, William Greenhalf, Andrew Owen, David G. Lalloo, Michael Jacobs, Julian A. Hiscox, Thomas Jaki, Pavel Mozgunov, Geoff Saunders, Gareth Griffiths, Saye H. Khoo, Tom E. Fletcher the AGILE CST-6 study group

## Abstract

**Background:** AGILE (NCT04746183) is a Phase Ib/IIa platform, evaluating candidates to treat COVID-19. CST-6 evaluated the safety and optimal dose of a novel intravenous (IV) formulation of favipiravir.

**Methods:** CST-6 was a dose-escalating, open-label, randomised, controlled, Bayesian adaptive Phase Ib trial. Hospitalised adults with PCR-confirmed SARS-CoV-2 infection, within 14 days of symptomatic COVID-19 were randomised 2:1 in groups of 6 (n = 4 favipiravir, n = 2 standard of care (SoC)) to ascending doses of IV favipiravir twice daily (b.i.d.) for 7 days or SoC. Clinical data, safety evaluations, virology and pharmacokinetic (PK) samples were collected. The primary outcome was safety. Secondary outcomes included clinical, PK and virological endpoints.

**Results:** 24 participants enrolled between 10/Sep/2022 and 01/Nov/2023 [10/24 female; median age 74 years (range 52-93)]. Favipiravir was well tolerated despite a high background rate of unrelated adverse events (AEs). No dose limiting toxicities (DLTs) were observed, with a model-predicted DLT risk of 16.8% and probability of unacceptable toxicity of 2.7% at the highest dose level. No SAEs were deemed related to favipiravir but an expected association with asymptomatic, transient hyperuricaemia was observed. PK exposures increased disproportionally to dose with significant accumulation in plasma, but with marked variability between participants within each cohort.

**Conclusions:** A novel formulation of favipiravir was safe at sustained high doses that reached PK targets in a study group with frailty and complex health profiles. Plasma concentrations demonstrated accumulation. Significant variability in PK parameters between individuals was noted. We consider doses up to 2400mg b.i.d. to be safe for further evaluation. https://clinicaltrials.gov/study/NCT04746183

**Key points:**

- A novel intravenous formulation of favipiravir, was safe and well tolerated in a frail and complex population, up to a dose of 2400mg b.i.d.
- Significant inter-individual variability in pharmacokinetic parameters was observed.
- Pharmacokinetic modelling suggests pre-specified target concentrations were met.

## Introduction

In the post-pandemic phase of COVID-19, novel directly-acting antiviral therapeutics are still required. AGILE (NCT04746183) is a multi-arm, multi-dose, phase Ib/IIa platform using a Bayesian adaptive design, established to address this need[1]. With a vaccinated population and the predominance of less pathogenic SARS-CoV-2 variants, patients hospitalised with symptomatic COVID-19 are now overwhelmingly frail, co-morbid or immunocompromised. These groups are often excluded from early phase clinical trials.

Favipiravir (6-fluoro-3-hydroxypyrazine-2-carboxamide; T-705) is an antiviral, licensed in oral form for use against influenza in Japan, with broad spectrum capabilities in non-clinical studies[2] that has been investigated in clinical studies of patients with influenza[3,4], Ebola virus disease [5] and COVID-19[6].

Favipiravir is rapidly and completely absorbed following oral administration[7]. Time to maximum plasma concentration is between 0.5 and 4.0 hours. Aldehyde oxidase (AO) is key in the metabolism of favipiravir; AO inhibition by favipiravir leads to accumulation of favipiravir in the plasma. Twice daily oral administration results in a greater than proportional rise in plasma concentration. Plasma protein binding averages 53-54%, 65% of which is due to albumin binding[7]. The major metabolite, T-705M1, is formed by AO in human liver cytosol and other tissues. The cytochrome mixed function oxidase systems do not play a significant role. Favipiravir and its metabolites T-705M1 and T-705M2 are excreted predominantly in urine, and to a lesser degree in faeces. No dose alterations are needed in renal impairment. Favipiravir is intra-cellularly phosphoribosylated to the active metabolite favipiravir ribofuranosyl-5’-triphosphate (T-705-RTP), which selectively inhibits RNA virus RNA-dependent RNA polymerases[8].

Uncertainty exists around the use of favipiravir as a treatment for COVID-19[6]. Concerns include risk of teratogenicity, pill burden and uncertain effectiveness against SARS-CoV-2[9–13]. *In vitro* studies demonstrate inconsistent activity against SARS-CoV-2, with EC_50_ values ranging from 61.88 µM to >500 µM, depending on experimental systems used[11,14–17]. Effective antiviral activity has been shown in animal models[18–20]. Clinical studies of oral favipiravir with pharmacokinetic (PK) characterisation show significant inter-individual variability in plasma concentrations[3,21] which in some cases diminish over time[4,22,23]. Differences in PK exposures may depend on weight, critical illness and ethnicity[3,22].

To maximise effectiveness, antiviral trough plasma concentrations should be maintained above the *in vitro* defined EC_90_ target[17,24]. The doses necessary to achieve this have been described by modelling based on published PK studies and likely concentrations of the active intracellular form (T-705-RTP)[25].

A novel intravenous (IV) formulation of favipiravir generates C_max_ levels 4-fold higher than oral dosing in cynomolgus monkeys[26,27], which may translate into sustained higher intracellular T-705-RTP concentrations and thus antiviral activity[25]. A 2021 single ascending dose (300-2400mg), healthy volunteer study reported no serious adverse events (SAEs)[28].

We sought to assess safety and tolerability of multiple ascending doses of IV favipiravir in participants hospitalised with COVID-19 and to recommend a safe dose for efficacy evaluation. Secondary objectives included PK characterisation and virological and clinical outcomes.

## Methods

### Study design, participants and ethics

This dose-escalation phase Ib study was designed as an open-label, randomised, controlled Bayesian adaptive trial in adult patients hospitalised with COVID-19, recruited in the NIHR Liverpool Clinical Research Facility (UK). Eligible participants were men and women aged ≥18 years with PCR-confirmed SARS-CoV-2 infection within 7 days of randomisation, within 14 days of symptom onset, with severity as defined by WHO Clinical Progression Scale grades 4-6. Women of childbearing potential (WOCBP) and men who were sexually active with WOCBP were required to use a highly effective method of contraception. The following criteria excluded participation: pregnant or breastfeeding, stage 4 or greater chronic kidney disease, alanine aminotransferase (ALT) and/or aspartate aminotransferase (AST) greater than 5 times the upper limit of normal, anticipated transfer to another hospital within 72 hours, known allergy to any study medication, participation in any clinical trial of an investigational medicinal product (CTIMP) within 30 days. All participants, or legally acceptable representatives, provided written informed consent before enrolment. The study protocol was reviewed by the UK Medicines and Healthcare products Regulatory Agency (MHRA) (EudraCT 2020–001860-27) and West Midlands Edgbaston Research Ethics Committee (20/WM/0136).

### Randomisation

Four sequential dosing tiers were defined *a priori* (600mg, 1200mg, 1800mg, 2400mg twice daily (b.i.d.) for 7 days or until discharge; a 300mg tier was allowed in case of de-escalation). Participants were recruited in cohorts of 6 and randomised to IV favipiravir: standard of care (SoC) in a 2:1 allocation ratio using permuted blocks with sentinel dosing in each cohort.

### Procedures

Inpatients at the Liverpool University Hospitals Foundation Trust with confirmed SARS-CoV-2 infection or symptoms consistent with COVID-19 were screened for eligibility. Baseline demographic and medical information was collected. Participants randomised to treatment received IV favipiravir twice daily for 7 days, or until discharge if this occurred before day 7. Clinical information and tests for safety monitoring, virology and PK were collected at predefined points. Additional safety tests were carried out if indicated.

Dose escalation was guided by the Bayesian adaptive design of the AGILE platform. As with previous candidates[29,30] a dose toxicity model[31] was used which estimates the probability of dose limiting toxicity (DLT, defined as any AE ≥ grade 3 CTCAE v.5, possibly or probably related to the IMP) at days 8 and 29 in control groups and at each dosing level[32]. A dose was deemed unsafe if the chance that treatment was associated with a 30% increase in the probability of a DLT occurring by day 8 was 25% or greater. An independent reviewer, blinded to allocation, adjudicated causality of all AEs ≥ grade 3, with those that were deemed possibly or probably related to treatment included into the model as DLTs. The Bayesian model recommended a course of action based on projected risk of DLT for the next dose level. On completion of each cohort, all available safety data were reviewed by the Safety Review Committee (SRC) who made the final decision on dose escalation, including ultimately a decision to recommend a safe dose for efficacy evaluation.

### Outcomes

Primary outcomes were DLTs up to day 8 and all AEs and SAEs in all participants. Secondary outcomes included plasma PK parameters up to day 8, change from baseline over time in viral load up to day 29, clinical progression at day 15 and 29 as quantified by the WHO Clinical Progression Scale, mortality at days 15 and 29 and time from randomisation to death (up to day 29).

### Pharmacokinetics

Plasma was sampled for favipiravir concentrations at days 1, 3 and 5. 2ml of venous blood was collected at 0-1 hours and 6-12 hours post-completion of infusion. Median T_last_ values for each sampling timepoint are described in table 3. On days 1 and 3 additional samples were taken pre-dose (C_0_ and C_trough_, respectively) and at 2-4 hours post-completion of infusion. Samples were cooled on wet ice, centrifuged (2000 g for 10 min at 4°C), distributed to 2 cryovials (approx. 500µL each) and transferred to a -80°C freezer. Drug concentrations were measured using a validated LC-MS/MS assay[33] at the Bioanalytical Facility (BAF) at the University of Liverpool.

Pharmacokinetic parameters were derived using non-compartmental analysis with Phoenix 64 WinNonlin (version 8.3).

### Pharmacokinetic Modelling

Plasma concentrations were modelled as a one-compartment[3,25] IV infusion population pharmacokinetic (pop-PK) model with first order clearance using nlmixr2 (version 3.0.2)[34]. Measurements below LLQ were labelled as 500 ng/mL with the exception of day 1 pre-dose measurements which were labelled as zero. A nonlinear mixed effects model was used to estimate the clearance and volume of distribution, scaled by bodyweight according to:

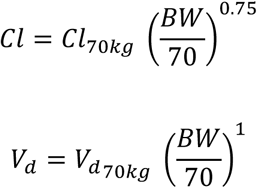

Where *Cl*_70*kg*_ and *V*_*d*_70*kg*__ denote the clearance and volume of distribution for an individual with a bodyweight of 70 kg respectively and *BW* is the bodyweight of the individual. A proportional error on plasma concentrations was also included. Prediction-corrected visual predictive check (PC-VPC) was produced using the built-in VPC function in nlmixr2.

For simulations, 2000 virtual participants were generated using Monte Carlo sampling of the bodyweight, clearance and volume of distribution, based on the probability distribution of fitted values. Bodyweights were sampled from a Gaussian distribution with the mean and standard deviation equal to those of the trial participants.

### Statistical analysis

All analyses are reported according to CONSORT 2010 and ICH E9 guidelines on Statistical Principles in Clinical Trials. All participants were included in both the evaluable population and the safety population for analysis. The primary endpoint of DLTs up to 8 days after first dose was modelled using a Bayesian dose–toxicity model based on Mozgunov et al[31,32]. The relationship between dose and toxicity was modelled using a two-parameter logistic model, where information can be shared across doses; the DLT rate in controls informs estimates for the active doses. The prior distributions for this model were calibrated to maximize the proportion of correct selection under a range of dose–toxicity scenarios where each dose considered was the optimum one. The toxicity risk in controls was *a priori* assumed to be 10%.

The model was updated after each cohort, with the final model presented as estimated DLT rates for each dose, alongside equal-tail 95% credible intervals. We report estimated additional toxicity above controls, the probability that the DLT rate falls within 15%–25% additional toxicity over controls (a predetermined acceptable range) and the probability of at least 30% additional toxicity over controls (deemed unacceptable).

Baseline demographics are summarised using descriptive statistics. Clinical endpoints are summarised at days 8, 15 and 29. The sample size was flexible, to adapt to cumulative safety data. Simulations were performed to assess model operating characteristics and to calibrate priors, assuming five doses (plus controls), with cohorts of size six capped at a total of 42 participants.

Statistical analysis was undertaken in SAS version 9.4, STATA version 16 and R version 3.6.0.

## Results

30 potential participants were identified for screening, 6 were excluded (WHO score, >14 days COVID-19 symptoms [x2], negative PCR, discharged before randomisation, unable to provide consent; described in figure 1). Baseline characteristics are described in table 1 and were similar across all groups. Four sequential dose cohorts (600, 1200, 1800 and 2400mg b.i.d.) of 6 participants each were randomised, and received doses of IV favipiravir or SoC in the period between 10/Sep/2022 and 01/Nov/2023. All participants randomised to treatment received at least 1 dose and 13 completed the full treatment course. 3 participants (1 in each of the 600, 1800 and 2400mg cohorts) did not complete the full course, due to discharge from hospital. One participant allocated to SoC requested to withdraw, prior to planned follow up at days 8, 11, 15 and 29.

**Figure 1.**
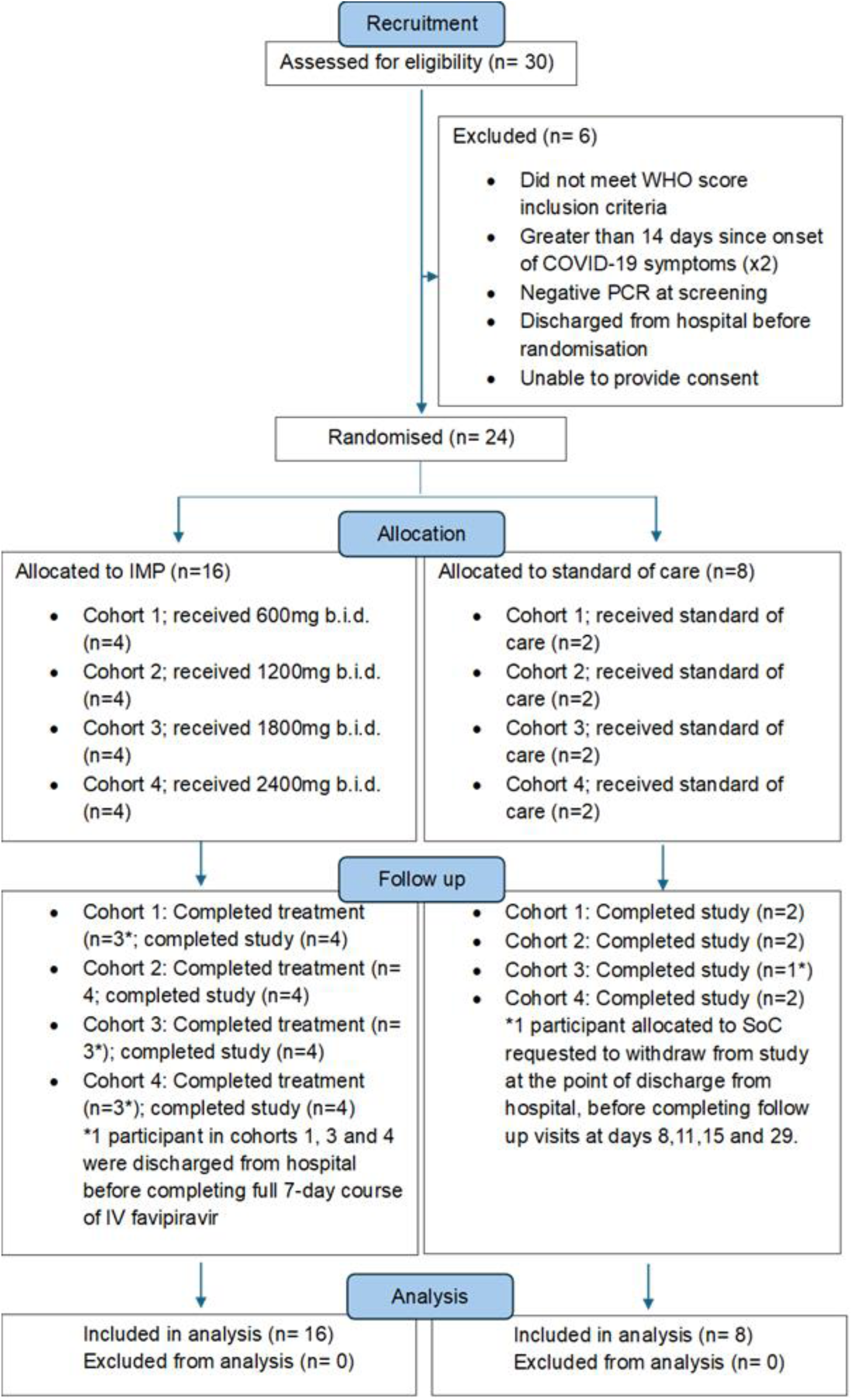
CONSORT diagram.

**Table 1.** Demographic and Baseline Characteristics – Evaluable Population.

| Demographic Characteristics | Favipiravir 600mg (N=4) | Favipiravir 1200mg (N=4) | Favipiravir 1800mg (N=4) | Favipiravir 2400mg (N=4) | Favipiravir Total (N=16) | Standard of Care (N=8) | Total (N=24) |
| --- | --- | --- | --- | --- | --- | --- | --- |
| **Age at consent (years)** |  |  |  |  |  |  |  |
| Median (Q1, Q3) | 82.5 (75.5, 90.5) | 76.5 (64.5, 84.0) | 66.0 (56.0, 79.0) | 76.5 (72.5, 81.5) | 76.5 (70.5, 86.0) | 73.5 (65.5, 77.0) | 74.0 (69.0, 81.0) |
| Min, Max | 74.0, 93.0 | 57.0, 87.0 | 52.0, 86.0 | 69.0, 86.0 | 52.0, 93.0 | 57.0, 81.0 | 52.0, 93.0 |
| **Gender, n (%)** |  |  |  |  |  |  |  |
| Male | 2.0 (50.0) | 2.0 (50.0) | 4.0 (100.0) | 1.0 (25.0) | 9.0 (56.3) | 5.0 (62.5) | 14.0 (58.3) |
| Female | 2.0 (50.0) | 2.0 (50.0) | 0.0 (0.0) | 3.0 (75.0) | 7.0 (43.8) | 3.0 (37.5) | 10.0 (41.7) |
| **Weight (kg)** |  |  |  |  |  |  |  |
| Median (Q1, Q3) | 68.3 (60.5, 78.1) | 70.80 (54.7, 96.3) | 85.4 (75.1, 107.0) | 87.1 (63.8, 99.5) | 78.6 (60.5, 93.9) | 78.45 (67.3, 102.9) | 78.45 (65.2, 99.3) |
| Min, Max | 57.2, 83.3 | 53.4, 107.0 | 68.4, 125.0 | 52.1, 100.1 | 52.1, 125.0 | 55.0, 117.4 | 52.1, 125.0 |
| **BMI (kg/m<sup>2</sup>)** |  |  |  |  |  |  |  |
| Median (Q1, Q3) | 25.8 (23.5, 27.2) | 27.0 (23.7, 32.3) | 28.2 (26.8, 35.9) | 30.0 (24.6, 36.8) | 27.8 (24.5, 30.2) | 25.5 (23.8, 32.9) | 27.2 (24.5, 31.0) |
| Min, Max | 22.3, 27.5 | 23.1, 34.9 | 25.4, 43.4 | 19.1, 42.7 | 19.1, 43.4 | 21.8, 36.2 | 19.1, 43.4 |
| **Ethnicity, n (%)** |  |  |  |  |  |  |  |
| White – English, Welsh, Scottish, Northern Irish | 4.0 (100.0) | 3.0 (75.0) | 4.0 (100.0) | 4.0 (100.0) | 15.0 (93.8) | 8.0 (100.0) | 23.0 (95.8) |
| White - other | 0.0 (0.0) | 1.0 (25.0) | 0.0 (0.0) | 0.0 (0.0) | 1.0 (6.3) | 0.0 (0.0) | 1.0 (4.2) |
| **First recorded COVID-19 infection** |  |  |  |  |  |  |  |
| Yes | 4.0 (100.0) | 3.0 (75.0) | 3.0 (75.0) | 3.0 (75.0) | 13.0 (81.3) | 7.0 (87.5) | 20.0 (83.3) |
| **Received vaccine for COVID-19** |  |  |  |  |  |  |  |
| Yes | 3.0 (75.0) | 4.0 (100.0) | 4.0 (100.0) | 3.0 (75.0) | 14.0 (87.5) | 6.0 (75.0) | 20.0 (83.3) |
| **WHO score (baseline)** |  |  |  |  |  |  |  |
| Median, range | 5.0 (5.0-5.0) | 4.0 (4.0-5.0) | 5.0 (4.0-5.0) | 5.0 (4.0-5.0) | 5.0 (4.0-5.0) | 5.0 (4.0-5.0) | 5.0 (4.0-5.0) |
| **Number of listed co-morbidities** |  |  |  |  |  |  |  |
| Median, range | 11.0 (9.0-13) | 11.5 (6.0-16.0) | 12.0 (5.0-19.0) | 11.0 (10.0-13.0) | 11.0 (5.0-19.0) | 10.5 (5.0-19.0) | 11.0 (5.0-19.0) |

N = Number of participants in the evaluable population.
BMI = body mass index
WHO score = World Health Organization clinical progression score

### Primary analysis

No DLTs were observed in any participant. Doses were escalated as planned to a maximum of 2400mg b.i.d. Bayesian model DLT point estimates with 95% credible intervals, calculated after each cohort, are shown in figure 2. The maximum dose level had an estimated DLT rate of 16.8% at day 8, with an estimated 11.2% additional toxicity over controls. The probability that toxicity fell within the predetermined target range of 15-25% additional toxicity over controls was 18.3%, which was the highest probability across all doses. The probability of ≥ 30% additional toxicity over controls was 2.7%. These values remained the same when data up to day 29 were included. These findings support the use of this dose in future phase II studies.

**Figure 2.**
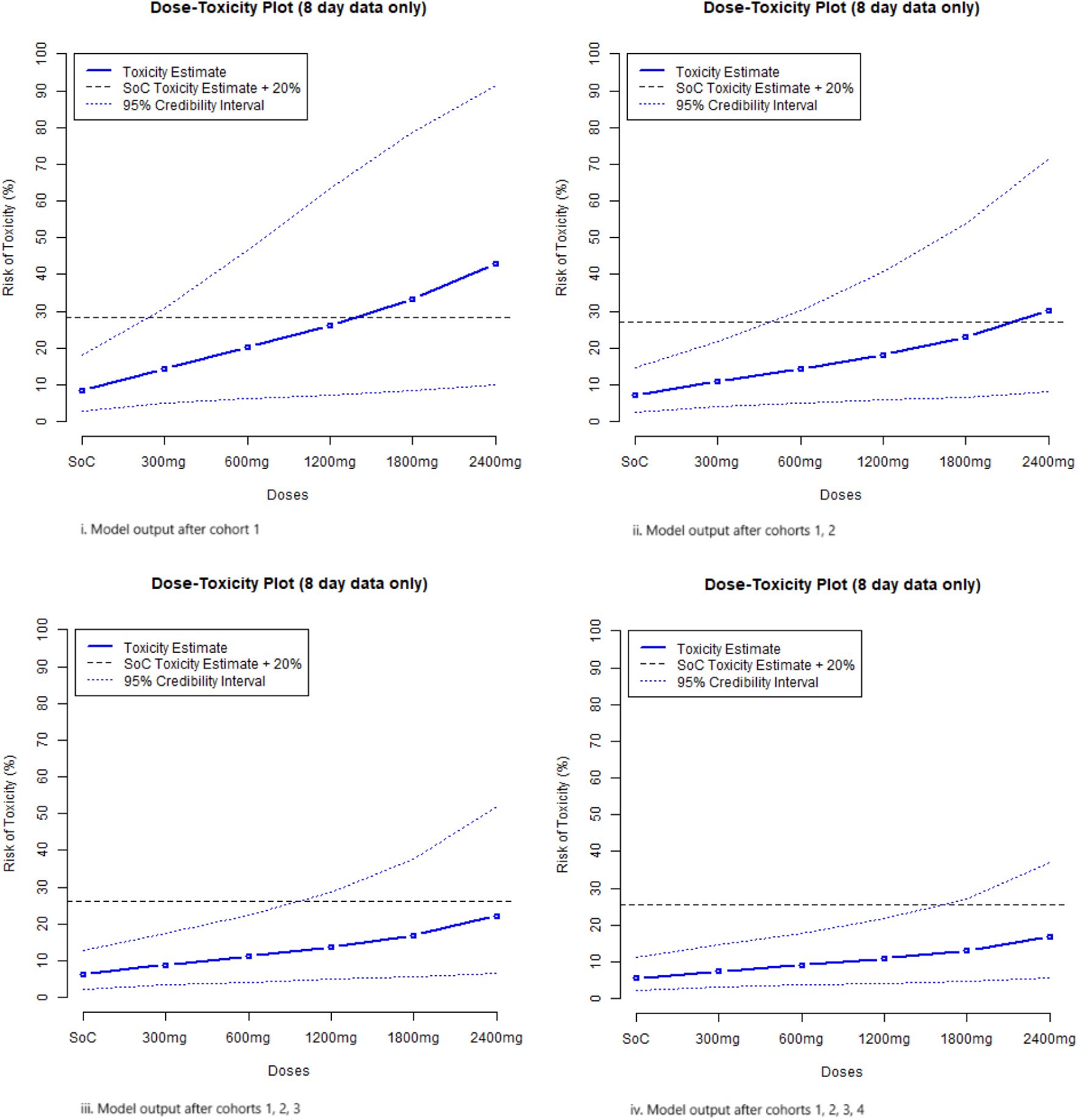
Bayesian model estimates of dose toxicity. Bayesian model output plots, updated after each cohort and presented as estimated DLT rates for each dose, alongside equal-tail 95% credible intervals. Plots remain identical after all data up to day 29 are included

AEs across all cohorts are detailed in table 2. At least 1 AE was experienced by all participants allocated to favipiravir. Of those allocated to SoC, 5 of 8 (62.5%) experienced at least 1 AE. AEs deemed related to favipiravir were experienced by 1 of 4 (25%), 3 of 4 (75%), 2 of 4 (50%) and 4 of 4 (100%) in the 600, 1200, 1800 and 2400mg b.i.d. cohorts respectively. All 10 related AEs were asymptomatic hyperuricaemia. The most common AEs by category were metabolism and nutrition disorders, infections and infestations and gastrointestinal disorders. AEs with severity ≥ grade 3 were experienced by 2 of 4 (50%), 2 of 4 (50%), 2 of 4 (50%) and 1 of 4 (25%) in the 600, 1200, 1800 and 2400mg b.i.d. cohorts respectively, and 4 of 8 (50%) of those allocated to SoC.

**Table 2.** Adverse Events by System Organ Class and CTCAE Term – Evaluable Population.

| System Organ Class | Favipiravir<br>600mg<br>(N=4)<br>n (%) | Favipiravir<br>1200mg<br>(N=4)<br>n (%) | Favipiravir<br>1800mg<br>(N=4)<br>n (%) | Favipiravir<br>2400mg<br>(N=4)<br>n (%) | Favipiravir<br>Total<br>(N=16)<br>n (%) | Standard<br>of Care<br>(N=8)<br>n (%) | Total<br>(N=24)<br>n (%) |
| --- | --- | --- | --- | --- | --- | --- | --- |
| **Participants with any System Organ Class** | 4.0 (100.0) | 4.0 (100.0) | 4.0 (100.0) | 4.0 (100.0) | 16.0 (100.0) | 5.0 (62.5) | 21.0 (87.5) |
| Blood and lymphatic system disorders | 1.0 (25.0) | 0.0 (0.0) | 0.0 (0.0) | 0.0 (0.0) | 1.0 (6.3) | 0.0 (0.0) | 1.0 (4.2) |
| Cardiac disorders | 0.0 (0.0) | 0.0 (0.0) | 0.0 (0.0) | 0.0 (0.0) | 0.0 (0.0) | 1.0 (12.5) | 1.0 (4.2) |
| Eye disorders | 0.0 (0.0) | 0.0 (0.0) | 1.0 (25.0) | 0.0 (0.0) | 1.0 (6.3) | 0.0 (0.0) | 1.0 (4.2) |
| Gastrointestinal disorders | 2.0 (50.0) | 0.0 (0.0) | 2.0 (50.0) | 2.0 (50.0) | 6.0 (37.5) | 1.0 (12.5) | 7.0 (29.2) |
| General disorders and administration site conditions | 3.0 (75.0) | 1.0 (25.0) | 0.0 (0.0) | 1.0 (25.0) | 5.0 (31.3) | 1.0 (12.5) | 6.0 (25.0) |
| Hepatobiliary disorders | 0.0 (0.0) | 1.0 (25.0) | 0.0 (0.0) | 0.0 (0.0) | 1.0 (6.3) | 0.0 (0.0) | 1.0 (4.2) |
| Immune system disorders | 0.0 (0.0) | 0.0 (0.0) | 0.0 (0.0) | 1.0 (25.0) | 1.0 (6.3) | 0.0 (0.0) | 1.0 (4.2) |
| Infections and infestations | 1.0 (25.0) | 1.0 (25.0) | 1.0 (25.0) | 3.0 (75.0) | 6.0 (37.5) | 3.0 (37.5) | 9.0 (37.5) |
| Injury, poisoning and procedural complications | 0.0 (0.0) | 0.0 (0.0) | 1.0 (25.0) | 1.0 (25.0) | 2.0 (12.5) | 0.0 (0.0) | 2.0 (8.3) |
| Investigations | 0.0 (0.0) | 1.0 (25.0) | 1.0 (25.0) | 0.0 (0.0) | 2.0 (12.5) | 1.0 (12.5) | 3.0 (12.5) |
| Metabolism and nutrition disorders | 3.0 (75.0) | 4.0 (100.0) | 2.0 (50.0) | 4.0 (100.0) | 13.0 (81.3) | 4.0 (50.0) | 17.0 (70.8) |
| Musculoskeletal and connective tissue disorders | 0.0 (0.0) | 0.0 (0.0) | 1.0 (25.0) | 1.0 (25.0) | 2.0 (12.5) | 1.0 (12.5) | 3.0 (12.5) |
| Nervous system disorders | 0.0 (0.0) | 1.0 (25.0) | 0.0 (0.0) | 3.0 (75.0) | 4.0 (25.0) | 0.0 (0.0) | 4.0 (16.7) |
| Psychiatric disorders | 2.0 (50.0) | 0.0 (0.0) | 0.0 (0.0) | 0.0 (0.0) | 2.0 (12.5) | 1.0 (12.5) | 3.0 (12.5) |
| Renal and urinary disorders | 1.0 (25.0) | 1.0 (25.0) | 0.0 (0.0) | 0.0 (0.0) | 2.0 (12.5) | 1.0 (12.5) | 3.0 (12.5) |
| Respiratory, thoracic and mediastinal disorders | 1.0 (25.0) | 1.0 (25.0) | 1.0 (25.0) | 2.0 (50.0) | 5.0 (31.3) | 2.0 (25.0) | 7.0 (29.2) |
| Skin and subcutaneous tissue disorders | 1.0 (25.0) | 1.0 (25.0) | 0.0 (0.0) | 0.0 (0.0) | 2.0 (12.5) | 0.0 (0.0) | 2.0 (8.3) |
| Vascular disorders | 0.0 (0.0) | 1.0 (25.0) | 0.0 (0.0) | 0.0 (0.0) | 1.0 (6.3) | 0.0 (0.0) | 1.0 (4.2) |
| N = Number of participants in the evaluable population.<br>Percentages are based on the total number of participants in each cohort. |  |  |  |  |  |  |  |

SAEs were experienced by 1 of 4 (25%), 1 of 4 (25%), 2 of 4 (50%) and 0 of 4 (0%) in the 600, 1200, 1800 and 2400mg b.i.d. cohorts respectively, and 1 of 8 (12.5%) of those allocated to SoC. No SAEs were deemed related to favipiravir.

### Analysis of secondary endpoints

### Clinical scores

There was no difference in the median WHO score between the treatment and SoC groups at baseline, day 8, day 15 and day 29. The 2400mg b.i.d. cohort had lower median WHO scores than the SoC group at days 8 and 15. Small numbers and clinical heterogeneity limit interpretation of these differences.

There were no deaths in all participants up to the end of the study.

### Pharmacokinetics

Significant variability in PK parameters between individuals within each cohort is the preeminent characteristic of these data, represented in figure 3. Plasma favipiravir was below the lower limit of quantification (LLQ; <1µg/mL) in all day 1 pre-dose samples, in all day 1 C_last_ (6-12hrs post completion of infusion, median T_last_ 6.1h) samples in the 600mg cohort, in all day 3 pre-dose samples and in 1 of 4 (25%) day 3 C_last_ samples in the 600mg cohort. In all other samples favipiravir was quantifiable, summary data are described in table 3. Plasma favipiravir concentrations demonstrated accumulation between day 1 and day 3 in all cohorts. At day 3, median favipiravir exposures (AUC_0-last_) for the 600mg (N = 3), 1200mg (N = 4), 1800mg (N=4) and 2400mg (N=4) cohorts were 38.54, 307.48, 383.03 and 1004.18 µg.h/mL respectively, with corresponding median C_max_ values of 16.80, 62.95, 87.51 and 200.99 µg/mL. Median day 3 C_trough_ values (pre-dose on day 3) were <1, 17.25, 19.46 and 96.49µg/mL. T_max_ was <1 hr in the 600, 1200 and 1800mg cohorts. In the 2400mg cohort T_max_ ranged from 0.08 to 6.28hrs. One plasma sample in the 600mg cohort on day 3 was excluded from all analyses due to a sampling error. A 1-compartment pop-PK model was produced using the data from this study (figure 4, 5). The estimated clearance and volume of distribution (table 4) conformed with previous studies of oral favipiravir[3]. Dose-stratified simulations show a mean C_trough_ exceeds the plasma target concentration of 159 μM at 2400 mg b.i.d. This suggests that a dose between 1800mg and 2400mg b.i.d. may be sufficient to achieve pre-specified PK targets for SARS-CoV-2[24].

**Figure 3.**
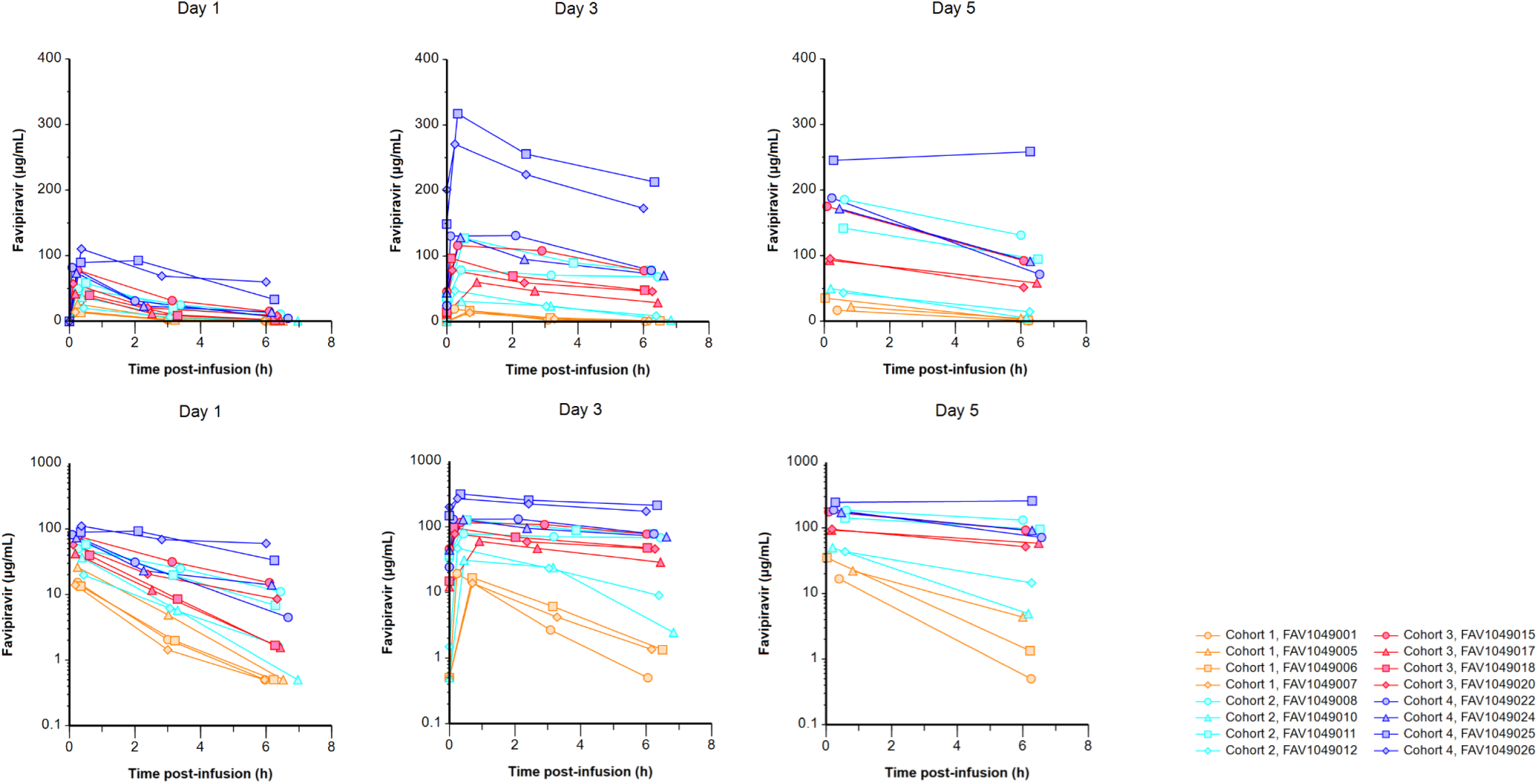
Favipiravir plasma pharmacokinetic profile. Favipiravir concentrations over actual time post-infusion stratified by study day in hospitalised patients with COVID-19 receiving intravenous favipiravir 600 mg twice daily over 1 hour (cohort 1: orange closed markers, solid lines), 1200 mg twice daily over 1 hour (cohort 2: cyan closed markers, solid lines), 1800 mg twice daily over 1 hour (cohort 3: red closed markers, solid lines) and 2400 mg over 1 hour (cohort 4: blue closed markers, solid lines) on a linear (top) and log-linear (bottom) scale (cohort 1: n=4 study Day 1, n=3 study Days 3 and 5; cohort 2: n=4 study Days 1, 3, and 5; cohort 3 and 4: n=4, study Days 1 and 3, n=3 study Day 5).

**Figure 4.**
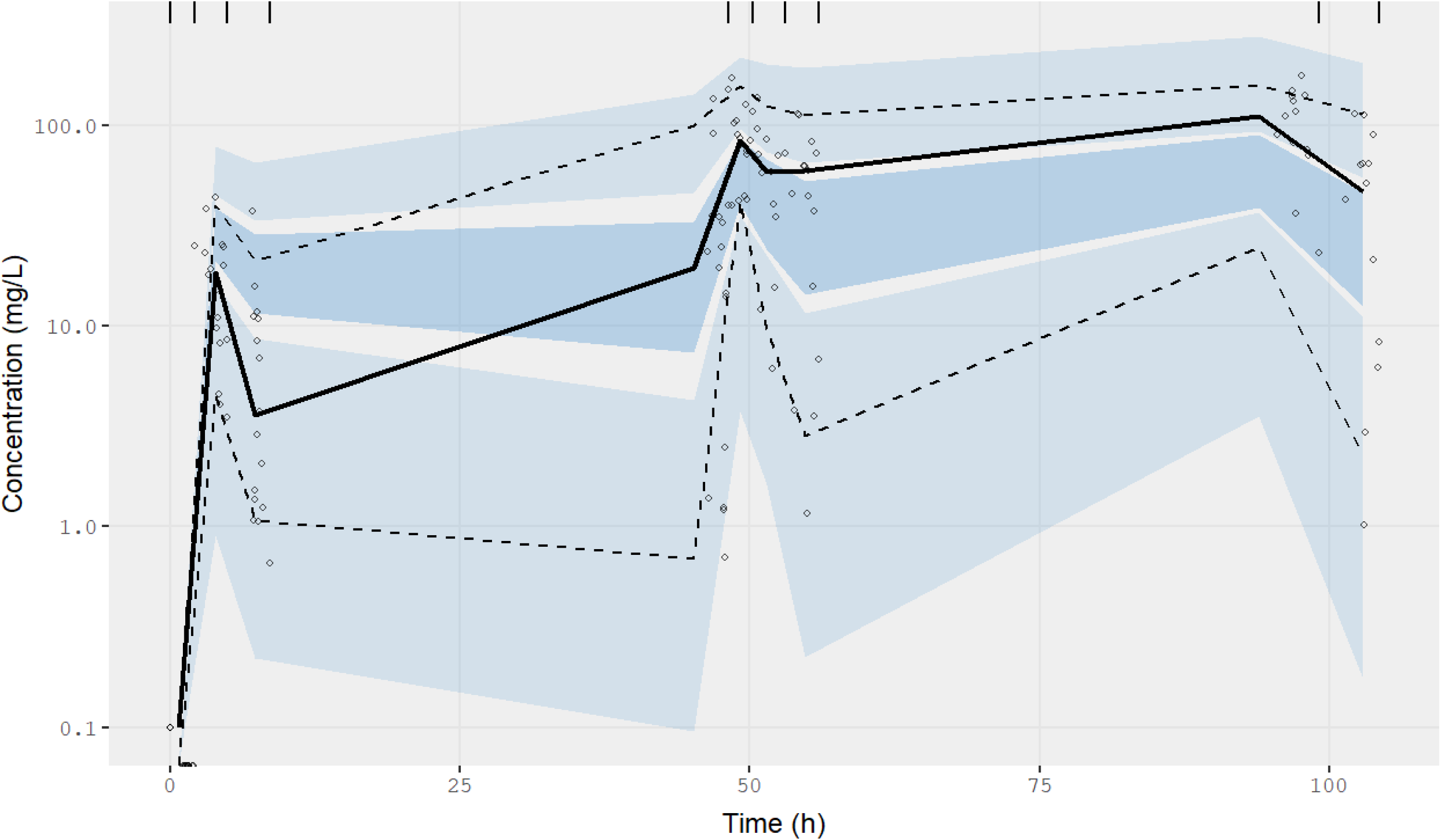
Population - PK modelling: Prediction-Corrected Visual Predictive Check (pcVPC). A pcVPC of the fitted 1-compartment IV infusion model. The solid black line represents the median and the dashed lines represent the 5^th^ and 95^th^ percentile of the data. The dark blue and light blue shaded areas represent the 90% prediction interval (5^th^ to 95 percentiles) of the simulated median, 5^th^ and 95^th^ percentiles based on 2000 simulated individuals. Binning for prediction intervals matched plasma measurement windows as indicated by vertical tick marks.

**Figure 5.**
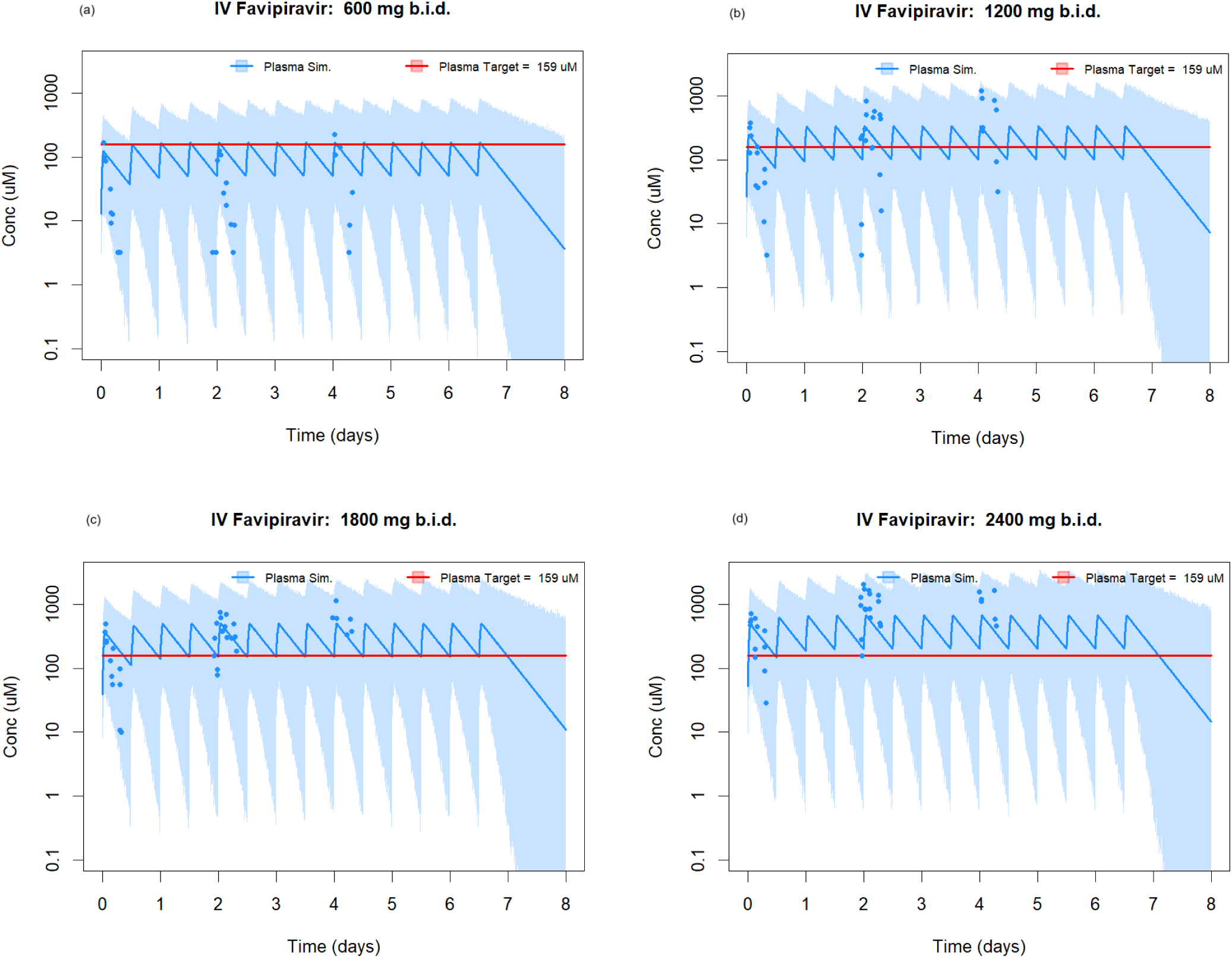
Population - PK modelling – simulations of plasma favipiravir concentrations. Simulations of favipiravir plasma concentrations following twice-daily, 1 hour IV infusions of favipiravir at 600 mg (a), 1200 mg (b), 1800 mg (c) and 2400 mg (d) b.i.d. The blue line and blue shaded area represent the median and the 90% prediction interval of 2000 simulations respectively. Observed data is represented by blue circles. The red line indicates the *in vitro* EC90 of favipiravir against SARS-CoV-2 in Vero-E6 cells[17]

**Table 3.** Summary of favipiravir plasma pharmacokinetic parameters in hospitalised patients receiving intravenous favipiravir.

| Day | Dose (mg, twice daily) | T<sub>max</sub> (h) | C<sub>0</sub>/trough (µg/mL)<sup>a</sup> | C<sub>max</sub> (µg/mL) | T<sub>last</sub> (h) | C<sub>last</sub> (µg/mL) | AUC<sub>0-last</sub> (h*µg/mL) |
| --- | --- | --- | --- | --- | --- | --- | --- |
| Day 1 | 600, N = 4 | 0.25 (0.18-0.35) | 0.00 (0.00-0.00) | 14.59 (13.34-26.03) | 6.10 (5.95-6.52) | 0.50<sup>a</sup> (0.50<sup>a</sup>-0.50<sup>a</sup>) | 28.74 (25.73-55.39) |
|  | 1200, N = 4 | 0.41 (0.27-0.50) | 0.00 (0.00-0.00) | 43.04 (19.83-58.21) | 6.41 (6.28-6.97) | 4.24 (0.50<sup>a</sup>-11.04) | 119.90 (51.45-177.80) |
|  | 1800, N = 4 | 0.22 (0.12-0.62) | 0.00 (0.00-0.00) | 49.55 (39.40-77.75) | 6.30 (6.10-6.43) | 5.11 (1.56-15.12) | 120.40 (91.68-236.20) |
|  | 2400, N = 4 | 0.29 (0.08-2.10) | 0.00 (0.00-0.00) | 87.17 (73.44-110.3) | 6.21 (6.00-6.67) | 23.57 (4.47-59.86) | 315.00 (179.00-444.50) |
| Day 3 | 600, N = 3 | 0.70 (0.23-0.70) | 0.50<sup>b</sup> (0.50<sup>b</sup>-0.50<sup>b</sup>) | 16.80 (13.83-19.42) | 6.17 (6.05-6.50) | 1.34 (0.50<sup>b</sup>-1.35) | 38.54 (36.31-46.68) |
|  | 1200, N = 4 | 0.44 (0.25-0.53) | 17.25 (0.50-37.10) | 62.95 (31.03-127.40) | 6.41 (6.20-6.83) | 38.73 (2.45-78.07) | 307.50 (130.20-601.60) |
|  | 1800, N = 4 | 0.25 (0.13-0.92) | 19.46 (12.15-46.17) | 87.51 (60.34-116.20) | 6.15 (6.02-6.43) | 47.00 (28.93-77.73) | 383.00 (270.30-605.10) |
|  | 2400, N = 4 | 0.38 (0.25-2.10) | 96.49 (24.46-201.40) | 201.00 (128.50-317.00) | 6.28 (6.00-6.62) | 125.50 (70.50-213.10) | 1004.20 (606.00-1592.30) |
| Day 5 | 600, N = 3 | 0.40 (0.03-0.82) | <sup>c</sup> | 22.10 (16.69-35.01) | 6.16 (6.00-6.25) | 1.34 (0.50<sup>a</sup>-4.35) | <sup>d</sup> |
|  | 1200, N = 4 | 0.58 (0.20-0.62) | <sup>c</sup> | 95.85 (43.25-185.50) | 6.24 (6.00-6.52) | 54.52 (4.92-131.20) | <sup>d</sup> |
|  | 1800, N = 3 | 0.17 (0.08-0.18) | <sup>c</sup> | 95.41 (92.89-175.10) | 6.22 (6.08-6.48) | 58.39 (51.50-92.31) | <sup>d</sup> |
|  | 2400, N = 3 | 0.47 (0.23-6.28) | <sup>c</sup> | 187.90 (171.40-258.50) | 6.38 (6.28-6.57) | 91.03 (71.52-258.50) | <sup>d</sup> |

All values represented as median (range)
T<sub>max</sub>: time after completion of infusion of maximum concentration
C<sub>0</sub>: pre-dose concentration, day 1
C<sub>trough</sub>: pre-dose concentration, day 3
C<sub>max</sub>: maximum measured concentration
T<sub>last</sub>: time after completion of infusion of last measured concentration
C<sub>last</sub>: concentration of the last measured timepoint
AUC<sub>0-last</sub>: area under the concentration-time curve from pre-dose to last measured sample
<sup>a</sup> sample prior to first dose of the day; Day 1 = C<sub>0</sub>, Day 3 = C<sub>trough</sub>

**Table 4.**
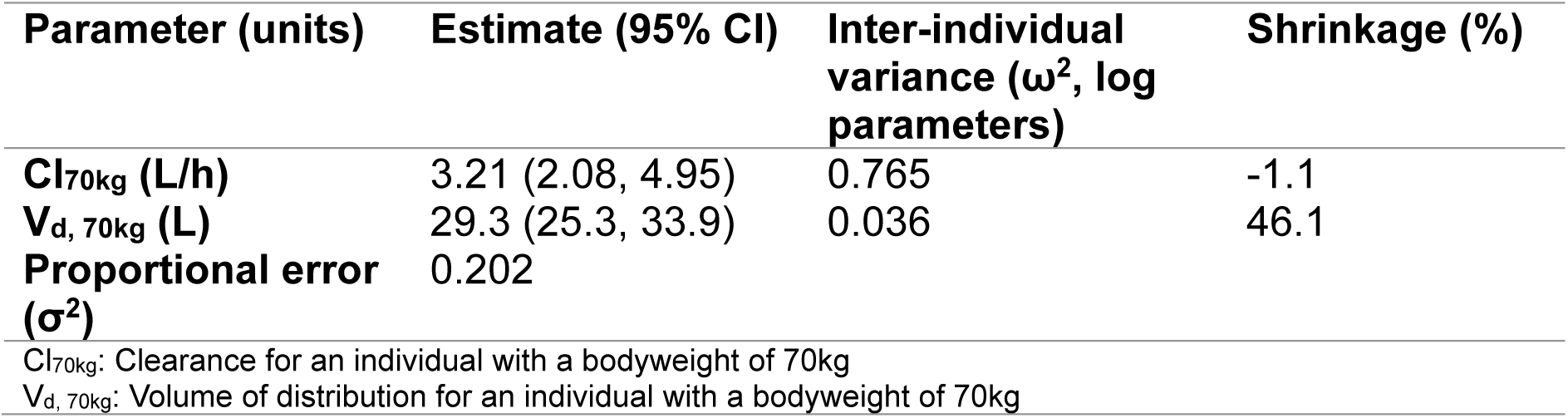
Summary of parameter estimates for favipiravir 1-compartment IV infusion POP-PK model fitting.

| Parameter (units) | Estimate (95% CI) | Inter-individual<br>variance ($\omega^2$, log<br>parameters) | Shrinkage (%) |
| --- | --- | --- | --- |
| **Cl<sub>70kg</sub> (L/h)** | 3.21 (2.08, 4.95) | 0.765 | -1.1 |
| **V<sub>d, 70kg</sub> (L)** | 29.3 (25.3, 33.9) | 0.036 | 46.1 |
| **Proportional error**<br>**($\sigma^2$)** | 0.202 |  |  |
| Cl<sub>70kg</sub>: Clearance for an individual with a bodyweight of 70kg |  |  |  |
| V<sub>d, 70kg</sub>: Volume of distribution for an individual with a bodyweight of 70kg |  |  |  |

### Virology

Detailed pharmacodynamic analysis is beyond the scope of this study. There was no significant difference in change from baseline viral load between the treatment and SoC groups. There was heterogeneity in individual drug exposure, concomitant antiviral medications and baseline viral load.

## Discussion

We describe the first use of IV favipiravir in hospitalised patients with SARS-CoV-2 infection and include characterisation of its safety, tolerability and pharmacokinetic profile. We demonstrate that IV favipiravir is safe and well tolerated at doses up to 2400mg b.i.d., in the context of the limited numbers of a phase Ib trial. AEs were reported in all cohorts, reflecting symptomatic COVID-19 and co-morbidity associated with a realistic patient population, characterised by co-morbidity and frailty to an unusual degree for early phase studies. A higher number of AEs was reported in participants allocated to treatment over SoC, however there was no dose dependent association. Hyperuricaemia was seen in 14 participants (10/16 allocated to favipiravir, 4/4 at the highest dose level; 4/8 allocated to SoC). In all cases this resolved following completion of treatment.

All AEs of severity ≥ grade 3 CTCAE v.5 were reviewed by a third-party independent assessor with access to all clinical data but blinded to treatment allocation; those deemed possibly or probably related to favipiravir were classified as DLTs and included in the model. In the final analysis the probability of greater than 30% excess toxicity over controls at 2400mg as estimated by the Bayesian model was 2.7%.

As with previous larger studies of oral favipiravir with PK characterisation [21], we observed marked inter-individual variability, supporting the idea of complex metabolism. The degree to which individual factors, including weight, sex, genetic variability, ethnicity and clinical status, contribute to this variability remains incompletely understood [3,22,35].

We considered that IV dosing may lead to transiently higher plasma concentrations, resulting in higher intracellular T-705-RTP levels[25], and thus greater activity and utility in more unwell patients. We did not use a loading dose, as our final target doses were in the range used as loading doses in other studies, and to investigate a previously observed decline in plasma favipiravir concentrations between days 3 and 5.

A significant fall in plasma favipiravir concentration has been observed after several days of oral dosing, following a loading dose and in critically unwell patients [4,23,36]. In our study accumulation in plasma was seen at day 3 and 5, suggesting differences in accumulation following a loading dose or potentially by route of administration. Our final cohort used higher sustained doses of favipiravir than previously published studies. In the 2400mg b.i.d. cohort, plasma favipiravir day 3 trough levels were close to or above the pre-specified target EC_90_ (24.9µg/mL; 159µM). Population-PK modelling suggests that doses of between 1800 and 2400mg b.i.d. may achieve pre-specified target concentrations (figures 4,5). Our data suggest that studies of oral favipiravir in COVID-19 using lower doses are unlikely to have achieved sustained plasma favipiravir concentrations above the target EC_90_. Indeed, a wide range of EC_50_ values have been reported and more recently both published and unpublished work suggest an EC_90_ closer to 500µM. Contrastingly, *in vivo* animal infection models have demonstrated potent antiviral effectiveness at considerably lower C_trough_ levels (mean C_trough_ 29.9μg/mL at the highest dose level) than those achieved in CST-6[18,20].

Interpretation of virological and clinical outcomes is limited by small numbers and heterogeneity of baseline data; this phase I study was not powered to assess efficacy of IV favipiravir against SARS-CoV-2.

IV favipiravir was well tolerated at doses up to 2400mg b.i.d. that reached pre-specified PK targets in a study group with frailty and complex health profiles. Our data support the safety of favipiravir at these doses in future phase II studies.

## Data Availability

The AGILE Trial Steering Committee will consider all reasonable requests by health-care providers, investigators, and researchers to provide anonymised data to address specific scientific or clinical objectives. The AGILE investigators are committed to reviewing requests from researchers for access to clinical trial protocols, de-identified patient-level clinical trial data, and study-level clinical trial data. Data will be assigned a DOI through deposition in the University of Liverpool Research Data Catalogue and shared under a Data Transfer agreement (or equivalent e.g. as part of a research collaboration agreement or confidentiality disclosure agreement.

## Acknowledgements

*The AGILE CST-6 study group

Shazaad Ahmad, Christopher J Edwards, Lesley Dry, Georgie McKenzie, Aleksandra Ros, Michael Stackpoole, Laura Bradley, Karen Jennings-Wilding, Nicholas Paton, Fred Hayden, Janet Darbyshire, Amy Lucas, Ulrika Lorch, Andrew Freedman, Richard Knight, Steven Julious

The supply of intravenous favipiravir was provided by Fujifilm Toyama as a gift, for which we would particularly like to acknowledge Koichi Yamada. We would like to thank to the staff of the Liverpool NIHR Clinical Research facility, and the staff at Liverpool University Hospitals Foundation Trust. We are deeply grateful to all participants for taking part in this trial.

## Author contributions

SHK, GG, RF, TF, TJ, TR, PM contributed to study design. SHK, GG, GS, WW, HP, AO, TF, TR, LD contributed to data analysis and interpretation. TF led clinical conduct as principal investigator of the clinical sites. RL, RF, LW, TR participated in clinical assessment and data collection. EC, LJE, VS, WG, CH, CK, KB, JAH contributed to study bioanalysis. KD, MI, YE, HER, MT, JG, JC, OO contributed to study management and execution. DGL, AO, MJ, TJ, PM contributed to the design of the AGILE platform. The manuscript was written by the authors, with TR as the overall lead author. No one who is not an author contributed to writing the manuscript. All authors had full access to the data; GS directly accessed and verified the underlying data reported in the manuscript relating to the Bayesian primary endpoint; KD directly accessed and verified all other data reported. The authors assume responsibility for the accuracy and completeness of the data and for the fidelity of the trial to the protocol.

## Data Sharing Statement

Alt Text

Figure 1. A CONSORT diagram graphically representing the disposition of participants within the trial.

Figure 2. Four charts, labelled i-iv, depicting the Bayesian model estimated dose-toxicity plots updated after each of the four cohorts, showing that the final estimate of toxicity falls below the pre-determined threshold for unacceptable toxicity.

Figure 3. Six plots of pharmacokinetic profiles, summarising individual participant plasma favipiravir over 0-6 hours on days 1, 3 and 5, with concentration (y-axis) first on a linear scale, then on a log scale

Figure 4. A prediction corrected visual predictive check plot of the fitted 1-compartment iv infusion model, with predicted favipiravir concentration plotted against time up to 100 hours

Figure 5. Four plots of the population-PK model outputs at each dose level, with simulated favipiravir concentrations plotted against time up to 7 days, showing that at the 2400mg b.i.d. dose, plasma target concentrations are exceeded throughout dosing

